# Minimal SARS-CoV-2 classroom transmission at a large urban university experiencing repeated into campus introduction

**DOI:** 10.1101/2022.03.16.22271983

**Authors:** Kayla Kuhfeldt, Jacquelyn Turcinovic, Madison Sullivan, Lena Landaverde, Lynn Doucette-Stamm, Davidson H. Hamer, Judy Platt, Catherine Klapperich, Hannah E. Landsberg, John H. Connor

## Abstract

SARS-CoV-2, the causative agent of COVID-19, has displayed person to person transmission in a variety of indoor situations. This potential for robust transmission has posed significant challenges to day-to-day activities of colleges and universities where indoor learning is a focus. Concerns about transmission in the classroom setting have been of concern for students, faculty and staff. With the simultaneous implementation of both non-pharmaceutical and pharmaceutical control measures meant to curb the spread of the disease, defining whether in-class instruction without any physical distancing is a risk for driving transmission is important. We examined the evidence for SARS-CoV-2 transmission on a large urban university campus that mandated vaccination and masking but was otherwise fully open without physical distancing during a time of ongoing transmission of SARS-CoV-2 both at the university and in the surrounding counties. Using weekly surveillance testing of all on-campus individuals and rapid contact tracing of individuals testing positive for the virus we found little evidence of in-class transmission. Of more than 140,000 in-person class events, only nine instances of potential in-class transmission were identified. When each of these events were further interrogated by whole-genome sequencing of all positive cases significant genetic distance was identified between all potential in-class transmission pairings, providing evidence that all individuals were infected outside of the classroom. These data suggest that under robust transmission abatement strategies, in-class instruction is not an appreciable source of disease transmission.

## Background

SARS-CoV-2, the causative agent of COVID-19, has posed significant challenges to day-to-day activities of colleges and universities. Many institutions of higher education faced immense challenges in 2020-2021 and again in 2021-2022, from spiking infections [1]. With the vaccination rollout, infection risk lessened, and in-class instruction largely resumed. To help limit SARS-CoV-2 transmission, some institutes of higher education instituted vaccination requirements for students and faculty along with non-pharmacological interventions such as surveillance testing, on-campus isolation of infected individuals, quarantine and frequent testing of close contacts, indoor masking, and enhancements to air filtration and circulation. These multi-faceted interventions allowed in-class instruction to fully resume and dormitories to reopen and re-populate. Despite these efforts, transmission has been continuously present in campus life.

Understanding whether in-class instruction without any physical distancing is fueling transmission is an important question that has not yet been adequately addressed. Multiple theoretical calculations have suggested different impacts of vaccination or masking on in-class transmission[2, 3], but real-world evaluation of classroom transmission is minimal. The lack of such data has prompted uncertainty among students, faculty, and staff regarding the infection risk of in-person instruction. To address this evidence gap, we sought to unambiguously identify instances of SARS-CoV-2 transmission in the classroom environment at a large urban university. Using a blend of adaptive surveillance testing, traditional epidemiology, and viral genomics, we analyzed incidence of likely transmission over more than 150,000 class meetings.

## Methods and Findings

We linked campus-wide surveillance testing, contact tracing, and class rosters (with permission under Boston University (BU) IRB 5693E) from 9/15/2021 to 12/1/2021. 9/1/2021 marked the full return to in-person instruction at BU. There were more than 150,000 in-person class meetings over this time period. Class sizes ranged from 2 to >400 individuals (average class size 31). Masking was mandatory for all students during in-person instruction. Faculty had the option to remove their mask while lecturing if they were >6 feet from the students and had increased their surveillance testing frequency. COVID-19 vaccination was mandated as of 9/2/2021, allowing for religious or medical exemption. Vaccination compliance at BU was defined as either completion of a SARS-CoV-2 vaccination series, documented medical, or religious exemption. Full vaccination rates for faculty, staff, and students respectively were 98.5%, 93.5%, and 98.7%.

SARS-CoV-2 transmission in the surrounding environment was significant over the study period (7 day rolling average in Suffolk County between 100-200 cases per day, in Middlesex: 200-600 per day). Boston University mandated regular weekly testing of the entire on-campus population [4]. During our analysis period of 8/1/2021 through November 31, 2021, more than 600,000 SARS-CoV-2 polymerase chain reaction (PCR) tests were conducted. Approximately 0.14% of these tests (896) showed detectable SARS-CoV-2 by reverse transcriptase real-time PCR (rRT-PCR) using the Centers for Disease Control and Prevention (CDC) primers for N1, N2[5].

All 896 instances of PCR positive samples were reported to the BU contact tracing team who initiated both direct personal interviews and class roster analysis to identify potential close contacts and sources of transmission. The overwhelming majority of cases investigated identified likely transmission events associated with living arrangements (e.g. roommates), socialization, or extended contact-time events. Of all PCR positive cases, a small number were deemed potential in-class transmission events. Potential instances of in-class transmission were defined as 2 or more SARS-CoV-2 positive individuals who shared an in-person class but did not identify one another as close contacts outside of the classroom environment. There were 9 instances of potential classroom transmission, accounting for approximately 0.0045% of all classroom meetings.

To more carefully investigate possible classroom transmission, we attempted whole virus genome sequencing from all SARS-CoV-2 positive samples identified during the study window. Sequencing was attempted using the Illumina COVIDSeq Assay [6] with a success rate of >90%. Full length genomes for each amplified sample were assembled through alignment to the Wuhan-Hu-1 reference sequence (NC_045512.2). Each genome was compared to itself and all other identified genomes. Comparing successfully sequenced samples during the study period and comparing genomic sequence relatedness with likely disease transmission scenarios identified through contact tracing, direct-transmission scenarios showed clear relatedness trends. Samples with zero nucleotide differences were overwhelmingly part of a linked transmission event. Genomes with 1 nucleotide difference were sometimes but not always related, and those with 2 or more nucleotide changes were considered highly unlikely to be transmission events. Genomes that differed by three or more nucleotides across 29,000 bases were considered to be not linked.

## Results

In analyzing potential classroom transmission events, seven of 9 potential transmission event pairs were identified as coming from different Pango lineages and as having >3 nucleotide differences. This made them unlikely to be classroom transmission events (Table 1). Of the remaining 2 potential classroom transmission events, instance 8 was illustrative of the low apparent in-class transmission. Three individuals who sat in close contact tested positive over a five-day span. Of those three, two had identical genomes upon sequencing. These two individuals were roommates (testing positive over a 6 day span) with significant unmasked exposure outside of the classroom. Living conditions were deemed the most likely cause of transmission. The individual sitting between these two roommates was diagnosed between when the two individuals tested positive (in the middle of the 6-day span noted above). Sequencing showed that this individual had a genome that differed by 2 nucleotide changes (Supplemental Figure 1), which is inconsistent with direct transmission from either individual in the classroom environment in the timeline presented.

**Table 1.** Instances of potential classroom-based transmission from August 1 to December 1, 2021.

| Potential Cluster Investigated | Date | # of Potential In-Class Transmission Individuals | # of Samples Sequenced | Genome Lineage | # of Nucleotide Differences between Samples | Additional Potential Exposure Points |
| --- | --- | --- | --- | --- | --- | --- |
| 1 | Early Sept | 2 | 2 | AY.103 & B.1.617.2 | 14 | Dining Halls |
| 2 | Early Sept | 2 | 2 | AY.3 & B.1.617.2 | 42 | Off-Campus Gathering, Dorm crossover |
| 3 | Early Sept | 2 | 1 | B.1.671.2 | Undetermined |  |
| 4 | Early Sept | 2 | 2 | AY.4 & B.1.617.2 | 40 | Boston Bars, Dental Clinic |
| 5 | Mid-October | 2 | 2 | AY.25 & B.1.617.2 | 22 | Dining Hall, Dorm Crossover |
| 6 | Late August | 2 | 2 | AY.4 & B.1.617.2 | 23 | Outside Boston Locations |
| 7 | Early October | 2 | 2 | AY.3 & AY.25 | 26 | Classroom building crossover |
| 8 | End Sept | 3 | 3 | AY.20 | $\leq 2$ | Public Transportation, Roommates, Lunch |
| 9 | Late November | 2 | 2 | AY.103 & AY.25 | 22 | Dining Hall, Building Overlap |

## Discussion

Our data support the hypothesis that a combination of SARS-CoV-2 vaccination and risk mitigation measures including indoor masking, regular surveillance testing, and enhanced air filtration can be highly effective at limiting disease spread within a large university academic environment to the extent that classroom transmission risk is negligible. With the ongoing concern of safety for students, faculty, and staff as universities return to fully normal functioning, this suggests an effective model for overall disease transmission safety in the classroom setting.

Limitations of our analysis include some inherent subjectivity in traditional epidemiologic investigations that center on an individual remembering all relevant interactions. This study occurred during a phase of the pandemic when the only variants circulating were associated with delta sublineages, and the findings may not apply to other SARS-CoV-2 variants. However, it does not at this point appear that the subsequent rise of the omicron variant increased the risk of in-class transmission. The omicron peak took place during a time when classes were not in session, and the return to class in 2022 was marked by rapidly falling disease incidence on campus despite in-person classes. This pattern is unlike the fall 2021 transmission pattern, where consistent transmission levels were seen over months of in-person classes.

## Data Availability

All data produced in the present study are available upon reasonable request to the authors

## Acknowledgements

Boston University provided financial support for the testing program described in this study and supported sequencing efforts. The BUMC Genome Sciences Institute provided partial financial support for sequencing. JHC acknowledges support from R21AI135517.

## Tables

**Supplemental Table 1.** Key statistics regarding classroom meetings, vaccination rates, non-pharmaceutical interventions, and SARS-CoV-2+ individuals detected on campus from August 1 to December 1, 2021. Abbreviations: eACH-effective air changes, Freq-frequency, Exch-exchanges.

| Time Period | # of Class Meetings | % Students Vaccinated | % Faculty Vaccinated | Testing Freq | Masking Policy | Typical Classroom Air Exch | SARS-CoV-2+ Individuals |
| --- | --- | --- | --- | --- | --- | --- | --- |
| Aug 1- Dec 1 | >150,000 | 98.7% | 98.8% | Once Weekly | Required In-Class | 4-6 eACH/h Merv13 minimum | 896 |

**Supplemental Figure 1.**
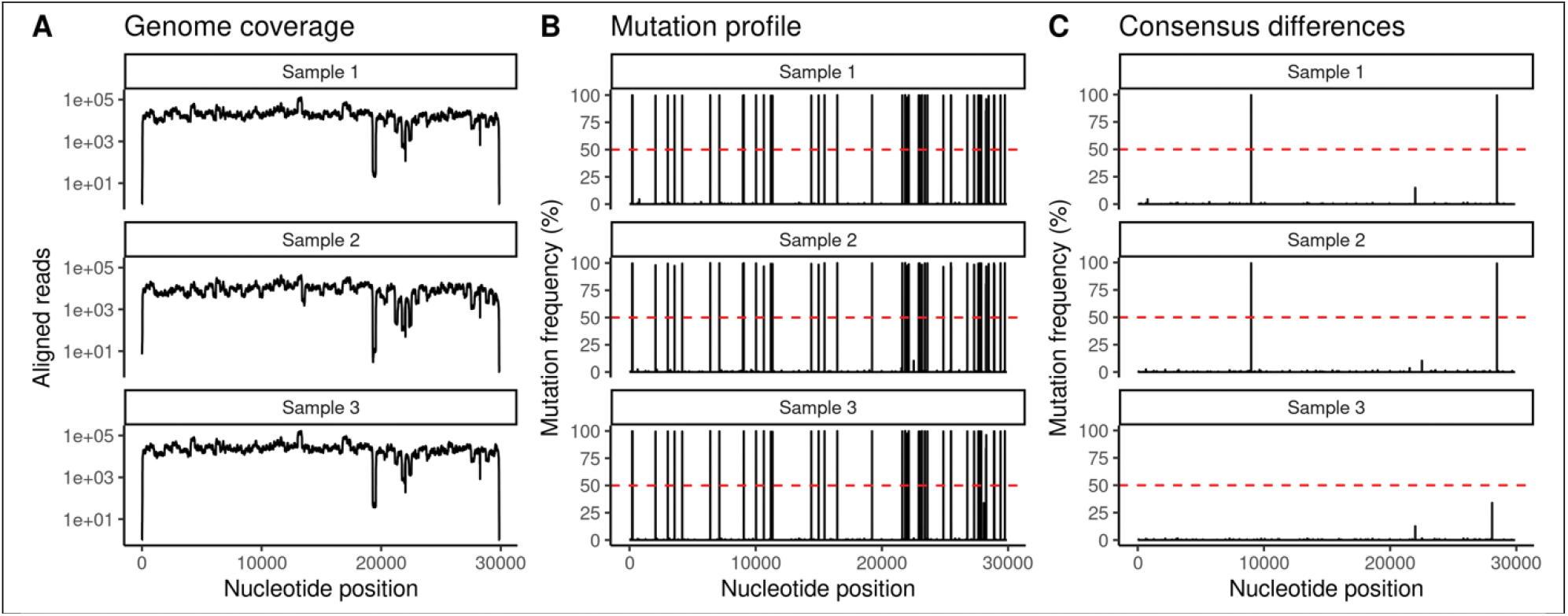
Mutation profiles of potential cluster 8. A) SARS-CoV-2 genome coverage plot showing read count at each SARS-CoV-2 nucleotide. All samples had >500X median coverage per sample B) SNV accumulation plots showing location of all accumulated variations in each sample. The full mutation profiles were very similar overall and showed no signs of contamination (dashed red line indicates 50% conversion from Wuhan NT at that position). C) SNV difference plots. Shared SNVs >50% were masked to highlight differences between samples (C). Samples 1 and 2 are identical; sample 3 lacks two nucleotide substitutions present at > 99% in samples 1 and 2 (8986 C > T and 28461 A > G).

